# Impact of COVID-19 restrictions on the postpartum experience of women living in Eastern Canada: A mixed method cohort study

**DOI:** 10.1101/2021.01.30.21250555

**Authors:** Justine Dol, Brianna Richardson, Megan Aston, Douglas Mcmillan, Gail Tomblin murphy, Marsha Campbell-yeo

**Affiliations:** Faculty of Health, Dalhousie University, Halifax, NS, Canada; School of Nursing, Faculty of Health, Dalhousie University, Halifax, NS, Canada; Nova Scotia Health Authority, Halifax, NS, Canada; Division of Neonatal Perinatal Medicine, Department of Pediatrics, Faculty of Medicine, Dalhousie University and IWK Health Centre, Halifax, NS, Canada

**Keywords:** postpartum adjustment, COVID-19, maternal self-efficacy, postpartum anxiety, postpartum depression

## Abstract

**Objectives:** To (1) compare changes in self-efficacy, social support, postpartum anxiety and postpartum depression in Canadian women collected before (Cohort 1) and during the COVID-19 pandemic (Cohort 2); (2) explore the women felt related to having a newborn during the pandemic; and (3) explore ways that women coped.

**Methods:** Prior to the pandemic (October 1, 2019-January 1, 2020), an online survey was conducted with women had given birth within the past six months in one of the three Eastern Canadian Maritime provinces (Cohort 1). A second, similar survey was conducted between August 1, 2020 and October 31, 2020 (Cohort 2) during a period of provincial pandemic response to COVID-19.

**Results:** For Cohort 1, 561 women completed the survey and 331 women in Cohort 2. Cohorts were similar in terms of age of women, parity, and age of newborn. There were no significant differences for self-efficacy, social support, postpartum anxiety, and depression between the cohorts. Difficulties that women reported as a result of COVID-19 restrictions included lack of support from family and friends, fear of COVID-19 exposure, feeling isolated and uncertain, negative impact on perinatal care experience, and hospital restrictions. Having support from partners and families, in-person/virtual support, as well as self-care and the low prevalence of COVID-19 during the summer of 2020 helped women cope.

**Conclusion:** While there was no significant difference in pre-pandemic and during pandemic psychosocial outcomes, there were still challenges and negative impacts that women identified. Consideration of vulnerable populations is important when making public health recommendations.

**What is already known on this subject?:** Previous work has shown the importance of social support in the postpartum transition in developing parenting self-efficacy and decreasing postpartum anxiety and depression. However, during the COVID-19 pandemic, women’s mental health, particularly during the perinatal period, has seen an increases in rates of postpartum anxiety and depression.

**What this study adds?:** This study is able to compare self-efficacy, social support, postpartum anxiety and depression between two cohorts of postpartum women living in Eastern Canada – pre-COVID-19 pandemic and during. While there was no significant difference in pre-pandemic and during pandemic psychosocial outcomes, there were still challenges and negative impacts that women identified.

---

The COVID-19 pandemic has disrupted many aspects of life. Women are disproportionally being impacted with significant impacts on childcare and their physical health (Connor et al., 2020). During the pandemic, there have been reports of increases in the rates of postpartum anxiety and depression (Davenport et al., 2020; Lebel et al., 2020; Ollivier et al., 2020; Wu et al., 2020). Previous work has shown the importance of social support in the postpartum transition in developing parenting self-efficacy (Leahy-Warren, 2005; Leahy-Warren et al., 2012) and decreasing postpartum anxiety and depression (Hetherington et al., 2018).

However, to minimize the spread of COVID-19, recommendations of physical distancing resulted in new mothers losing that social connection, isolating at home without any in-person support – both from healthcare providers and family and friends. There is currently limited knowledge on the impact that COVID-19 has had on the postpartum experience of new mothers, particularly as compared to pre-pandemic level.

The objectives of this study were to (1) compare changes in self-efficacy, social support, postpartum anxiety and postpartum depression in Canadian women collected before (Cohort 1) and during the COVID-19 pandemic (Cohort 2); (2) explore the women felt related to having a newborn during the pandemic; and (3) explore ways that women coped.

## Method

This is a mixed methods cohort study. Prior to the pandemic (October 1, 2019-January 1, 2020), a survey was conducted with women living in the Maritime provinces (Cohort 1) as part of another research project. Following the first wave of COVID-19 in Canada, a similar survey was conducted between August 1 and October 31, 2020 (Cohort 2). Ethical approval was granted by the IWK Health Research Ethics Board (#1024525)

### Participants

Women were considered eligible if they: (1) had an infant six months of age or less; (2) were over 18 years of age; (3) were able to speak, write and read English; and (4) currently live in a Maritime Province. Women were excluded if they lived outside the Maritime Provinces or had their most recent baby more than six-months prior to the date the survey was completed.

### Setting

The study was completed across three Eastern Canadian provinces, Nova Scotia, New Brunswick and Prince Edward Island, often referred to as the Maritime provinces. They make up 5.1% of Canada’s population (Statistics Canada, 2019) and 4.2% of Canadian births (Statistics Canada, n.d.). States of emergency were declared between March 11 and 22, 2020 across the three provinces. On July 3, 2020, all three provinces with Newfoundland and Labourer created the ‘Atlantic Bubble’, which kept open borders between provinces without having to self-isolate upon arrival (The Council of Atlantic Premiers, 2020). Individuals arriving from elsewhere in Canada or internationally were still required to self-isolate for 14 days. The Atlantic bubble remained until November 24^th^, 2020 when self-isolation was again required in some provinces to minimize the impact of the second wave of the pandemic.

In terms of epidemiology of COVID-19, Nova Scotia experienced their first wave from approximately March to May 2020, with very few cases from June until October 1, which was deemed the start of their second wave (Government of Nova Scotia, 2020). New Brunswick’s first wave was approximately March to June 2020, with a lull in the summer and the second wave starting October 2020 (New Brunswick, n.d.). Prince Edward Island had only 89 total cases March to December (Government of Prince Edward Island, 2020).

Throughout the pandemic, there have been significant restrictions in all three provinces including: required self-isolation of symptomatic individuals, non-essential travel restrictions between provinces or region, limits on social gatherings, masks required in public places, and physical distancing requirements. In specific relation to the perinatal experience, there were restrictions on the number of support people allowed to be present during antenatal appointments, during childbirth, and for postpartum appointments – either being no support people allowed or only one support person. There was a reduction or cancellation of in-person healthcare visits from public health nurses, parent support groups, and public health drop-ins.

### Procedure

Women were recruited to participate in an online survey through online advertisements (e.g., Facebook, Twitter), media outreach, and study posters. For both surveys, women could opt into a draw for one of three $100 CAN electronic gift cards. Once participants completed the eligibility screening, they were directed to a consent form. The survey contained standardized measures and open ended response options which took approximately 30 minutes to complete.

### Measures

Self-efficacy was measured using the Karitane Parenting Confidence Scale (KPCS), a 15 item scale which assesses perceived self-efficacy of mothers of newborns birth to twelve months (Crncec et al., 2008). A cut off score of 39 or less (out of a possible 45) is considered clinically low perceived parenting self-efficacy (Crncec et al., 2008). Social support was measured through the Multidimensional Scale of Perceived Social Support (MSPSS), a 12-item scale to measure how much support a parent feels they get from family, friends and significant others from which

subscales from each can be calculated (out of 4) as well as a total score (out of 12) (Zimet et al., 1990). Postpartum anxiety was measured using the Postpartum Specific Anxiety Scale (PSAS), a 51-item scale which evaluates anxiety specifically relevant to the postpartum period (Fallon et al., 2016). A score of 112 out of 204 is considered clinical levels of anxiety, with higher scores indicating greater anxiety (Fallon et al., 2016). Postpartum depression was measured using the Edinburgh Postnatal Depression Scale (EPDS)(Cox et al., 1987)(Cox et al., 1987)(Cox et al., 1987), a 10-item self-report screening scale that indicates if a respondent has symptoms related to postnatal depression, with a score of 14 or greater indicates ‘probable depression’ (Cox et al., 1987, 2014).

To determine the impact of COVID-19 on the postpartum experience, participants in Cohort 2 were asked to share the hardest part about having a newborn during COVID-19 and what helped them cope. Participants also completed COVID-19 exposure and impact questions and the Impact of Stressful Event Scale – Revised (IES-R), a 22-item self-report assessment of subjective distress caused by traumatic events with scores ranging from 0 (low distress) through 12 (high distress) (Weiss, 2007).

### Data Analysis

Independent sample t-tests were used to determine differences between Cohort 1 and Cohort 2 on self-efficacy, social support, postpartum anxiety, and depression. Potential differences across parity and age of newborn were also considered. Means and standard deviations were used to describe the sample. Open ended responses were analyzed by the first author and verified by the second using thematic analysis informed by a qualitative descriptive approach (Bradshaw et al., 2017; Braun & Clarke, 2006).

## Results

### Participants

For Cohort 1, the survey was completed by 561 women who had given birth between April to December 2019. For Cohort 2, the survey was completed by 331 women who had given birth between February to October 2020. The women were similar in terms of parity, geographic location, age of newborn, and number of biological children with demographic details in Table 1.

**Table 1.** Demographics of cohorts.

| **Demographics** | **Cohort 1**<br>**n=561**<br>**n (%)** | **Cohort 2**<br>**n=331**<br>**n (%)** |
| --- | --- | --- |
| Infant Age |  |  |
| 0-3 months | 309 (55.1) | 214 (64.7) |
| 4-6 months | 252 (44.9) | 117 (35.3) |
| Parity |  |  |
| Primiparous | 317 (56.5) | 213 (64.4) |
| Multiparous | 244 (43.5) | 118 (35.6) |
| Province |  |  |
| Nova Scotia | 261 (46.5) | 221 (66.8) |
| New Brunswick | 171 (30.5) | 75 (22.7) |
| Prince Edward Island | 129 (23.0) | 35 (10.6) |
| Marital Status |  |  |
| Single | 12 (2.1) | 8 (2.4) |
| In a relationship | 67 (12.0) | 23 (6.9) |
| Married/common-law | 479 (85.4) | 298 (90.1) |
| Missing | 3 (0.5) | 2 (0.6) |
| Household Income (CAN) |  |  |
| Less than \$49,000* | 100 (17.8) | 48 (14.5) |
| \$50,000-\$149,000 | 356 (63.5) | 208 (62.9) |
| Over \$150,000 | 74 (13.2) | 60 (18.1) |
| Missing | 31 (5.5) | 15 (4.5) |
| Baby birth weight |  |  |
| Under 2,500 grams | 30 (5.4) | 13 (3.9) |
| 2,500-4,000 grams | 360 (64.2) | 232 (70.1) |
| Over 4,000 grams | 96 (17.1) | 49 (14.8) |
| Missing | 75 (13.3) | 37 (11.1) |
| Birth Method |  |  |
| Vaginal birth | 407 (72.5) | 228 (68.9) |
| Unplanned cesarean | 102 (18.2) | 68 (20.1) |
| Planned cesarean | 52 (9.3) | 34 (10.3) |
| Prefer not to answer | - | 1 (0.3) |

\* For pre-COVID, this was spilt by under \$45,000 and between \$45,000-\$149,000

### Comparing psychosocial outcomes pre- and during pandemic

As shown in Table 2, there were no significant differences on any of the psychosocial outcomes between Cohorts. In Cohort 1, 31.9% of the sample had clinical high anxiety symptoms (PSAS>112) and 20.3% had high depression (EPDS>14) compared to 31.9% and 18.0% of Cohort 2, respectively. When considering differences by parity, the only significant difference was for primiparous women with social support from friends, which was lower in Cohort 1 (M=5.54, SD=1.42) than Cohort 2 (M=5.80, SD=1.29), p=0.033. When considering age of their baby, the only significant difference was again with social support from friends (cohort 1 M=5.34, SD=1.50; cohort 2 M=5.73, SD=1.34, p=0.014) and total social support (cohort 1 M=5.77, SD=1.09; cohort 2 M=6.00, SD=0.96, p=0.039) for women with infants aged 4-6 months. No other differences were found (non-significant results not shown).

**Table 2.** Psychosocial outcomes measured pre and during COVID-19 pandemic.

| **Outcome** | **Cohort 1**<br>**Mean (SD)** | **Cohort 2**<br>**Mean (SD)** | **p value** |
| --- | --- | --- | --- |
| Self-efficacy | 37.29 (5.21) | 37.27 (5.46) | 0.957 |
| Social Support<br>(Family) | 5.82 (1.29) | 5.83 (1.37) | 0.789 |
| Social Support<br>(Friends) | 5.50 (1.45) | 5.67 (1.36) | 0.069 |
| Social Support<br>(Significant Other) | 6.38 (0.98) | 6.44 (0.91) | 0.320 |
| Social Support<br>(Overall) | 5.90 (1.02) | 5.98 (1.00) | 0.238 |
| Postpartum anxiety | 99.98 (26.32) | 102.11 (26.97) | 0.292 |
| Postpartum depression | 9.27 (5.32) | 8.76 (5.08) | 0.153 |

### COVID-19 Stressors

Of the 331 respondents in Cohort 2, 3.3% were potentially exposed to someone with COVID-19 and 5.1% were suspected to have COVID-19. No one had a positive test. Twelve percent (12.4%) said their relationship with family members got a lot or a little better, 19.3% said it got a lot or little worse, and 66.8% had no change. In terms of financial problems, 24.7% said COVID-19 created moderate to extreme problems whereas 75.2% indicated slight or no financial impact. Just over half (53.5%) said their personal contacts decreased, 36.6% said contacts increased, and 8.5% stayed about the same. In terms of the IES-R, the mean was 0.81 (SD = 0.72, range = 0.00-3.64).

### Negative impact of COVID-19 on postpartum experience

Women mentioned several difficulties they experienced as a result of COVID-19 restrictions, including lack of support from family and friends, fear of COVID-19 exposure, feeling isolated and uncertainty, negative impact on perinatal care experience, and hospital restrictions.

One of the biggest difficulties was the lack of support available from their families and friends in the immediate postpartum period. One woman explained that this is “the one time in your life you should have family and friends around and we weren’t allowed.” Similarly, women discussed how hard it was for them not to have their families and friends meet their newborn in person, particularly if they lived in different provinces. A woman explained “My husband is an essential worker and had to return to work after the baby was born and no one could visit because they lived in another province. For 2 months I was completely alone with my baby”.

Women also indicated their fear around COVID-19 exposure for their newborn, themselves, or their other family members. Women were afraid to get groceries or go to appointments in case they were exposed to the virus and brought it back home. One woman explained: “[I] worry about bringing COVID home to my child. Anytime I need to go get groceries or something with my baby with me, I worry that I’m putting him at risk for selfish reasons.” Even if women had visitors, women discussed the difficulties of having people visit while maintaining safety protocols and distance for their newborn and their families. For example, one woman said “knowing that I need to be very selective about who holds my newborn but feeling a lot of pressure by friends and family to allow them to hug/hold/kiss him. I feel very judged and like I am being an overbearing mother if I set boundaries…”

Women also reported feeling very isolated and alone during the postpartum time. Women discussed the “being alone for months”, the “isolation from friends and family”, and the feeling of being “a bit trapped and isolated”. They also felt very uncertain about what has happening or going to happen: “uncertainties about how life would be affected by COVID” and “stress of the unknown”.

Women also discussed the negative impact of COVID-19 on their perinatal care experience. Women were disappointed and often struggled with having to attend antenatal appointments alone, without their partner or a support person. One woman explained: “This was my first baby, and my fiancé was unable to attend half of the appointments/ultrasounds that we had. He lost out on important moments.” This resulted in not all women receiving their desired antenatal care and not being able to attend in-person prenatal classes. Similarly, women mentioned not having sufficient postpartum care, with virtual care often not living up to what they needed. One woman said: “I had my newborn in March when everything was closing, I felt like I had less support because everything was over the phone, all my in-person postpartum appointments were canceled.” Women also commented on their lack of postpartum peer support groups and missing out on the postpartum experience they had expected. For example, women felt “like my maternity leave has been ripped away”, the “lack of mom-groups for that peer-to-peer support,” and “the feeling of missing out on the normal experiences of being a new parent.”

Women mentioned the challenges due to hospital rules and restrictions which negatively influenced their postpartum experience as there was much uncertainty around who could be present when they were going to deliver. “[I] worried about giving birth in the hospital with the regulations constantly changing and not allowing or limiting support people.” Women also commented that they ended up labouring alone or did not have their desired delivery support.

One woman said: “I gave birth alone because my partner had a cold and wasn’t allowed in the hospital”. Another woman said “My husband [was] not allowed in the hospital until I was about to deliver. I was there 22 hours by myself (this was my first child) - a very lonely and scary ordeal!”

### Coping with COVID-19 during the postpartum period

Women also reflected on what helped them cope during this challenging period and acknowledged that this came from having support from partners, support from families, in-person/virtual support, as well as engaging in self-care and the low epidemiology of COVID-19 during the summer of 2020.Women reported the importance of partner support to help them cope during COVID-19 restrictions. One woman said: “My husband [has helped me cope]. Him and I against the world!” Some women indicated their partner was supportive and helpful in household responsibilities. Other women commented on the benefit of having their partner work from home or take parental leave and the extra bonding that allowed as a family. One woman explained: “Our first baby was born end of April and it was a unique experience getting to have that uninterrupted time together to really bond with our baby… Also, he’s been working from home during COVID, so he’s really been able to be more present than he would have normally and that has been so nice for the both of us.”

Women reported that support from their family was essential. The amount of support varied, depending on the restrictions with women expressing relief when some restrictions on household numbers and inter provincial travel were lifted during the summer months. One woman wrote that “allowing my mom to come help us and be part of our bubble” was important in helping her cope, while another said, “My family was very supportive and made many physical distanced doorstop visits and left us meals.” Some women also commented on the essential nature of having support from families, even when it was against the public health recommendations. A woman explained that “having my mom stay with us for the first two weeks despite that being prohibited” helped her cope while another said she “broke the ‘rules’ and had [her] mother come and help out after a week of exhaustion.”

Women mentioned support from friends and community members as important to helping them cope, including talking to friends, having people drop off meals, and having support from people in their ‘bubbles’. Women indicated the importance of virtual connection with family and friends via FaceTime or text message: “FaceTime! Thank goodness for FaceTime so at least my family could easily see the baby.” Similarly, women also mentioned having support from online sources, such as online mom groups. One woman said: “Reading and participating in online forums and new mom groups made me feel less alone and was a great way to ask questions and share experiences.”

Beyond support, women also mentioned the importance of engaging in self-care, such as going for walks or exercising, as well as focusing on their newborn and their new family was helpful during this time. One woman explained: “I enjoyed the first few months of being able to bond with my baby and husband and create a routine/stay at home.” Women also commented on their focus of this experience being a time-limited concern and that things will get better: “Just taking things day by day, trying not to watch the news and just reminding myself that my baby is safe with me and that eventually this will all be a memory.”

Finally, the low COVID-19 prevalence in the Maritimes was reassuring to women. One woman said: “Living in a rural area, where there have been no cases reported helped. We also physical distanced and I self-isolated with the baby until he was one month old.” Another woman commented on the importance of how lifting restrictions helped: “Being able to have our “bubble” opened up and have my parents and in-laws visit and help with older children.”

## Discussion

This study explored the impact of COVID-19 on the postpartum experience of women living in the Maritime provinces. While there was no significant difference in pre-pandemic and during pandemic on psychosocial outcomes, there were still important challenges and negative impacts that women identified. In particular, the lack of support from family and friends, fear of COVID-19 exposure, feeling isolated, hospital restrictions and uncertainty negatively impacted on perinatal care experience. Having support from partners, families, and in-person/virtual support was helpful for women in coping with the impacts of COVID-19.

It is reassuring that there were no significant differences in self-efficacy, social support, postpartum anxiety and depression between cohorts. This differs than previous work which found an increase in perinatal anxiety and depression due to the COVID-19 pandemic (Lebel et al., 2020). However, no significant differences were found in a meta-analysis in eight studies for postpartum depression but there was an increase in anxiety (Hessami et al., 2020). In our sample, not only was the epidemiology in the area extremely low, the percentage of the population that was exposed to, or suspected to having, COVID-19, was also low. This may have played a role in decreasing the influence that COVID-19 had on maternal mental health. Additionally, the support women received from their partners, family and friends may have provided some resilience and buffer against the more extreme impacts found in other studies to date.

It is clear that women still felt negatively impacted by the restrictions related to COVID-19. The impact of lockdowns and physical restrictions has been shown to result in negative mental health and well-being outcomes, suggesting that while physical distancing recommendations can reduce the spread of COVID-19, it is not without cost to psychological well-being (Every-Palmer et al., 2020; White & Van Der Boor, 2020). In other perinatal populations around the world, similar feelings around the lack of support, fear of COVID-19 exposure, feeling isolated and uncertain, negative impact on perinatal care experience, and hospital restrictions were significant challenges that women faced during this period (Farewell et al., 2020; KC et al., 2020; Nanjundaswamy et al., 2020; Pant et al., 2020).

The “bubbling” with families and the larger Atlantic bubble seemed to alleviate some difficulties, with increased access to support from family and friends. In a study in New Zealand adults, who had a similar low number of cases yet also experienced a period of lockdown, they similarly found positive, ‘silver linings’ including family time and ability to focus on self-care and pausing and reflecting (Every-Palmer et al., 2020). Other studies have found that partners are an essential component in helping cope with the challenges with having a newborn during the COVID-19 pandemic and providing resiliency (Farewell et al., 2020). Thus, considerations of this vulnerable population are essential in planning physical distancing recommendations. Consideration of two-household families may be especially important for families who are experiencing a life transition, such as welcoming a newborn, but may also be recommended for those suffering from illness or other health concerns. It is important to understand how different coping methods can be utilized to reduce the negative health risks of isolation and lockdowns.

## Limitations

While this study had several strengths, there were some limitations. This study was only conducted in the Maritime provinces, which had a low prevalence of COVID-19 and is not necessarily presentative of the whole Canadian experience. Additionally, while the sample size was considerable for the qualitative responses, it may not have been sufficiently powered to detect differences on the psychosocial outcomes. Thus, these findings should be interpreted in light of these limitations.

## Conclusion

While there were no significant differences between pre-pandemic and during pandemic psychosocial outcomes, there were still important challenges and negative impacts women identified related to their postpartum experience. The lack of support from family and friends, fear of COVID-19 exposure, feeling isolated and uncertain, negative impact on perinatal care experience, and hospital restrictions negatively impacted women. While this was buffered through means of coping, including support from partners and families as well as self-care, consideration of vulnerable populations is important when making public health recommendations for society.

## Data Availability

Data is not available for sharing due to confidentiality of participant response.

## References

Bradshaw, C., Atkinson, S., & Doody, O. (2017). Employing a Qualitative Description Approach in Health Care Research. Global Qualitative Nursing Research, 4. https://doi.org/10.1177/2333393617742282

Braun, V., & Clarke, V. (2006). Using thematic analysis in psychology. Qualitative Research in Psychology, 3(2), 77–101. https://doi.org/dx.doi.org/10.1191/1478088706qp063oa

Connor, J., Madhavan, S., Mokashi, M., Amanuel, H., Johnson, N. R., Pace, L. E., & Bartz, D. (2020). Health risks and outcomes that disproportionately affect women during the Covid-19 pandemic: A review. Social Science & MedicineSocial, 266(113364).

Cox, J., Holden, J., & Henshaw, C. (2014). The Edinburgh Post Natal Depression Scale (EPDS) Manual.

Cox, J., Holden, J., & Sagovksy, R. (1987). Detection of Postnatal Depression: Development of the 10-item Edinburgh Postnatal Depression Scale. British Journal of Psychiatry, 150, 782–786. https://doi.org/10.1007/978-94-007-1694-0_2

Crncec, R., Barnett, B., & Matthey, S. (2008). Development of an instrument to assess perceived self-efficacy in the parents of infants. Research in Nursing and Health, 31(5), 442–453. https://doi.org/10.1002/nur.20271

Davenport, M., Meyer, S., Meah, V. L., Strynadka, M. C., & Khurana, R. (2020). Moms are not OK: COVID-19 and maternal mental health. Frontiers in Global Women’s Health, 1(June), 1–6. https://doi.org/10.3389/FGWH.2020.00001

Every-Palmer, S., Jenkins, M., Gendall, P., Hoek, J., Beaglehole, B., Bell, C., Williman, J., Rapsey, C., & Stanley, J. (2020). Psychological distress, anxiety, family violence, suicidality, and wellbeing in New Zealand during the COVID-19 lockdown: A cross-sectional study. PLoS ONE, 15(11 November), 1–19. https://doi.org/10.1371/journal.pone.0241658

Fallon, V., Halford, J. C. G., Bennett, K. M., & Harrold, J. A. (2016). The Postpartum Specific Anxiety Scale: development and preliminary validation. Archives of Women’s Mental Health, 19(6), 1079–1090. https://doi.org/10.1007/s00737-016-0658-9

Farewell, C. V., Jewell, J., Walls, J., & Leiferman, J. A. (2020). A Mixed-Methods Pilot Study of Perinatal Risk and Resilience During COVID-19. Journal of Primary Care and Community Health, 11. https://doi.org/10.1177/2150132720944074

Government of Nova Scotia, C. (2020). Coronavirus (COVID-19): case data. Webpage. https://novascotia.ca/coronavirus/data/

Government of Prince Edward Island. (2020). PEI COVID-19 Case Data. Webpage. https://www.princeedwardisland.ca/en/information/health-and-wellness/pei-covid-19-case-data

Hessami, K., Romanelli, C., Chiurazzi, M., & Cozzolino, M. (2020). COVID-19 pandemic and maternal mental health: a systematic review and meta-analysis. Journal of Maternal-Fetal and Neonatal Medicine, 0(0), 1–8. https://doi.org/10.1080/14767058.2020.1843155

Hetherington, E., McDonald, S., Williamson, T., Patten, S. B., & Tough, S. C. (2018). Social support and maternal mental health at 4 months and 1 year postpartum: analysis from the All Our Families cohort. Journal of Epidemiology and Community Health, 72, 933–939. https://doi.org/10.1136/jech-2017-210274

KC, A., Gurung, R., Kinney, M. V., Sunny, A. K., Moinuddin, M., Basnet, O., Paudel, P., Bhattarai, P., Subedi, K., Shrestha, M. P., Lawn, J. E., & Målqvist, M. (2020). Effect of the COVID-19 pandemic response on intrapartum care, stillbirth, and neonatal mortality outcomes in Nepal: a prospective observational study. The Lancet Global Health, 8(10), e1273–e1281. https://doi.org/10.1016/S2214-109X(20)30345-4

Leahy-Warren, P. (2005). First-time mothers: Social support and confidence in infant care. Issues and Innovations in Nursing Practice, 50(5), 479–488. https://doi.org/10.1111/j.1365-2648.2005.03425.x

Leahy-Warren, P., Mccarthy, G., & Corcoran, P. (2012). First-time mothers: Social support, maternal parental self-efficacy and postnatal depression. Journal of Clinical Nursing, 21(3–4), 388–397. https://doi.org/10.1111/j.1365-2702.2011.03701.x

Lebel, C., MacKinnon, A., Bagshawe, M., Tomfohr-Madsen, L., & Giesbrecht, G. (2020). Elevated depression and anxiety among pregnant individuals during the COVID-19 pandemic. Journal of Affective Disorders, 277, 5–13. https://doi.org/10.31234/osf.io/gdhkt

Nanjundaswamy, M. H., Shiva, L., Desai, G., Ganjekar, S., Kishore, T., Ram, U., Satyanarayana, V., Thippeswamy, H., & Chandra, P. S. (2020). COVID-19-related anxiety and concerns expressed by pregnant and postpartum women—a survey among obstetricians. Archives of Women’s Mental Health. https://doi.org/10.1007/s00737-020-01060-w

New Brunswick. (n.d.). New Brunswick COVID-19 Dashboard. Webpage. Retrieved December 16, 2020, from https://experience.arcgis.com/experience/8eeb9a2052d641c996dba5de8f25a8aa

Ollivier, R., Aston, D. M., Price, D. S., Sim, D. M., Benoit, D. B., Joy, D. P., Iduye, D., & Nassaji, N. A. (2020). Mental Health & Parental Concerns during COVID-19: The Experiences of New Mothers Amidst Social Isolation. Midwifery, 102902. https://doi.org/10.1016/j.midw.2020.102902

Pant, S., Koirala, S., & Subedi, M. (2020). Access to Maternal Health Services during COVID-19. Europasian Journal of Medical Sciences, 2(2), 48–52. https://doi.org/10.46405/ejms.v2i2.110

Statistics Canada. (n.d.). Live births, by place of residence of mother. 2020. Retrieved June 10, 2020, from https://www150.statcan.gc.ca/t1/tbl1/en/tv.action?pid=1310041401&pickMembers%5B0%5D=2.8

Statistics Canada. (2019). Annual demographic estimates: Canada, provinces and territories. https://www150.statcan.gc.ca/n1/pub/91-215-x/91-215-x2019001-eng.htm

The Council of Atlantic Premiers. (2020). Atlantic Provinces Form Travel Bubble. In News release. https://immediac.blob.core.windows.net/cap-cmha/images/Newsroom/Draftnewsrelease(v7).pdf

Weiss, D.. (2007). The Impact of Event Scale-Revised. In J. P. Wilson & T. M. Keane (Eds.), Assessingpsychological trauma and PTSD: a practitioner’s handbook (2nd Editio, pp. 168–189). Guilford Press.

White, R. G., & Van Der Boor, C. (2020). Impact of the COVID-19 pandemic and initial period of lockdown on the mental health and well-being of adults in the UK. BJPsych Open, 6(5), 1–4. https://doi.org/10.1192/bjo.2020.79

Wu, Y., Zhang, C., Liu, H., Duan, C., Li, Y., Fan, J., Li, H., Chen, L., Xu, H., Li, X., Guo, Y., Wang, Y., Li, X., Li, J., Zhang, T., You, Y., Li, H., Yang, S., Tao, X.,… Huang, H. (2020). Perinatal depressive and anxiety symptoms of pregnant women along with COVID-19 outbreak in China. American Journal of Obstetrics and Gynecology.

Zimet, G. D., Powell, S. S., Farley, G. K., Werkman, S., & Berkoff, K. A. (1990). Psychometric Characteristics of the Multidimensional Scale of Perceived Social Support. Journal of Personality Assessment, 55(3–4), 610–617. https://doi.org/10.1080/00223891.1990.9674095

